# Comorbidities and sociodemographic factors on COVID-19 fatalities

**DOI:** 10.1101/2022.05.20.22275397

**Authors:** Jacob Gerken, Demi Zapata, Daniel Kuivinen, Isain Zapata

## Abstract

**Introduction:** Previous studies have evaluated comorbidities and sociodemographic factors individually or by type but not comprehensively. This study aims to analyze the influence of a wide variety of factors in a single study to better understand the big picture of their effects on case-fatalities.

**Methods:** County-level comorbidities, social determinants of health such as income and race, measures of preventive healthcare, age, education level, average household size, population density, and political voting patterns were all evaluated on a national and regional basis. Analysis was performed through Generalized Additive Models and adjusted by CCVI.

**Results:** Factors associated with reducing COVID-19 case fatality rates were mostly sociodemographic factors such as age, education and income, and preventive health measures. Obesity, minimal leisurely activity, binge drinking, and higher rates of individuals taking high blood pressure medication were associated with increased case fatality rate in a county. Political leaning influences case case-fatality rates. Regional trends showed contrasting effects where larger household size was protective in the Midwest, yet harmful in Northeast. Notably, higher rates of respiratory comorbidities such as asthma and COPD diagnosis were associated with reduced case-fatality rates in the Northeast. Increased rates of CKD within counties were often the strongest predictor of increased case-fatality rates for several regions.

**Conclusion:** Our findings highlight the importance of considering the full context when evaluating contributing factors to case-fatality rates. The spectrum of factors identified in this study must be analyzed in the context of one another and not in isolation.

## INTRODUCTION

The SARS-CoV-2 Virus, also known as the COVID-19 virus, has currently led to over 990,000 (April, 2022) deaths in the United States ^1^. The virus has ravaged not only the United States financially and put a stop to every in-person activity for the last two years ^2^ but has highlighted how embedded community traits affect the outcome of a whole community. In the end, it is characteristics like comorbidities and sociodemographic factors that play in a defining role in determining a community’s fatality outcome ^3^.

While it has been shown that a wide range of comorbidities has an impact COVID-19 outcomes; there has been a great effort to define the specific contributions of comorbidities in their impact on COVID-19 morbidity and mortality rates ^4–7^. Comorbidities such as hypertension ^8^, diabetes mellitus ^9^, chronic kidney disease ^10^, chronic obstructive pulmonary disease ^11^, and cardiovascular disease ^12^ among others, have important repercussions on COVID-19 outcomes. However, there are also a variety of reasons for which to consider some sociodemographic factors as deleterious, such as lacking healthcare insurance, being of a specific racial background ^7,13^, or even voting patterns. All these factors combined would point in a direction that suggests that differences in COVID-19 fatality rates are a potential outcome of embedded community characteristics.

Even though previous studies have examined factors influencing COVID-19 fatalities ^4–6,14^, no study has performed a comprehensive analysis of all the aforementioned factors together to the same extent as our study. In our study, a multitude of key community indicators such as comorbidities, sociodemographic factors (including voting patterns), and determinants of health have been examined to reflect trends and potential associations that can be compared against each other. Therefore, the objective of this study was to perform a comprehensive evaluation of comorbidities, sociodemographic factors, and determinants of health at a national level using county aggregated to define their association to COVID-19 case-fatalities. These patterns may allow us to alter the way communities handle public health crises, utilize public health interventions that could deflect harmful outcomes, and provide resources to communities in a timely manner based on their community characteristics.

## METHODS

### Datasets

The focus of the study was to evaluate regional trends of COVID-19 case-fatality rate compared to comorbidities and sociodemographic factors. This study was vetted and categorized as exempt by the Institutional Research Board. Our study utilized countywide data for each county in the entire continental United States and Hawaii; Alaska and Puerto Rico were excluded from the analysis due to differences in their county data reporting. COVID-19 case-fatality rates were gathered from the COVID-19 Community Profile Report ^15^ for the January 2-8, 2021 week cutoff, this report included the CCVI ^16^. This cutoff week was selected because it allowed for the evaluation of the COVID-19 case fatality rate, without the influence of the vaccines or newer, more infectious strains. Rates of various comorbidities such as chronic obstructive pulmonary disease (COPD), hypertension, cancer, asthma, chronic heart disease (CHD), cholesterol, diabetes, chronic kidney disease (CKD), smoking, stroke, and obesity were obtained on a per-county basis from the CDC 2020 Population Level Analysis and Community Estimates (PLACES) project ^17^. Rates of poor mental health, binge drinking, lack of health insurance, time allocated to leisurely activity, and preventive care consisting of cervical cancer screening, routine doctor visits, dental visits, cholesterol screening, and routine mammography were obtained from the 2020 CDC PLACES Project, as well. Other variables such as average household size and population density for each county were acquired from the United States Census Bureau COVID-19 website ^18^. Latitude of each county was also included in the analysis and obtained from the United States Gazetteer Files ^19^ from the United States Census Bureau. The 2020 Presidential voting records of each county were obtained from the Harvard Dataverse ^20^. Racial makeup in each county was obtained from the 2020 decennial United States census ^21^, while income, age, and education level were retrieved from the 2019 American Community Survey 5-year estimates ^22^.

### Statistical analysis

Data was evaluated for associations using a Generalized Additive Models (GAMs) approach. COVID-19 case fatality rates per 100k people were set as the dependent variable while each comorbidity, sociodemographic and health determinant factor was set as an independent variable. All models were adjusted using COVID-19 Community Vulnerability Indexes (CCVI) ^16^ which normalized the data for inherent inequity ^23^. This study uses individual counties as the experimental unit. All analyses were evaluated on the national sample and in a per region samples, as to determine regional patterns. Independent variables were introduced into the model using smoothing splines starting with three degrees of freedom. Models assumed Gaussian residual distributions. All analyses were performed using PROC GAM in SAS/STAT v.9.4 (SAS Inc., Cary, NC). Risk ratios were estimated with 99% confidence intervals and the coefficients sign determined effect directions. Negative coefficients indicated a reduction in COVID-19 case-fatality rates while positive coefficients indicated an increase in case-fatalities. Coefficient standardization was done with a normally distributed Z-score transformation. All associations presented were tested using two-tailed tests. Regional pattern models were performed independently to identify the top contributors - negatively and positively associated. Even with 99% confidence intervals, all tests were declared significant at a Bonferroni threshold. Regional pattern top contributors that did not reach the Bonferroni threshold are indicated in the figures.

## RESULTS

### National level trends

Data from 3140 counties was included in our analysis. Our GAM approach examined the data for associations to case-fatalities, all these values were adjusted to CCVI to normalize the inherent differences a raw risk ratio would estimate, and the risk ratio standardized estimates are presented in Figure 1. Raw estimates showcase the extent of the association without considering their spread while standardized estimates adjust the extent of the association to the spread. Raw estimate findings (Figure 1A) revealed that comorbidities above sociodemographic factors have the largest effects associated with case-fatalities; however, these associations can go in both positive and negative directions. A diagnosis of cancer provided the largest effect decreasing COVID-19 case-fatalities while CKD and stroke had the largest effects increasing them. Similarly, asthma (decreased risk), CHD (increased risk) and diabetes (increased risk) displayed moderate effects. Household size was the largest significant sociodemographic factor in positive outcomes of COVID-19 with an effect in a range comparable to relevant comorbidities. Other demographic effects such as age and education displayed significant associations that reduced or increased case-fatality risk. Populations with higher educational achievements displayed significantly reduced case-fatality rates. Increased income always displayed a protective effect. Political preference was significantly associated with case-fatalities such as voting for Biden reduced case-fatalities while voting for Trump had the opposite effect. Racial and ethnic backgrounds were only associated to COVID-19 case–fatalities for Pacific Islanders, Asian and Black groups. Determinants of health such as cervical cancer screening and people using high blood pressure medication also showed mixed direction associations. Cervical cancer screening had the largest case-fatality reducing effect from this category while the people using high blood pressure medication had the largest opposite effect. Standardized estimates (Figure 1B) show a different perspective and allow for comparisons across factors as they are standardized. In this case, a routine colonoscopy procedure was found to be the largest protecting effect against COVID-19 case-fatality followed by a combination of sociodemographic factors such as age, education, and income. On the other side of the spectrum, obesity had the largest negative impact deleterious outcome in COVID-19 patients followed by having no leisure physical activity, binge drinking and higher proportions of people taking high blood pressure medication in a specific county.

**Figure 1.**
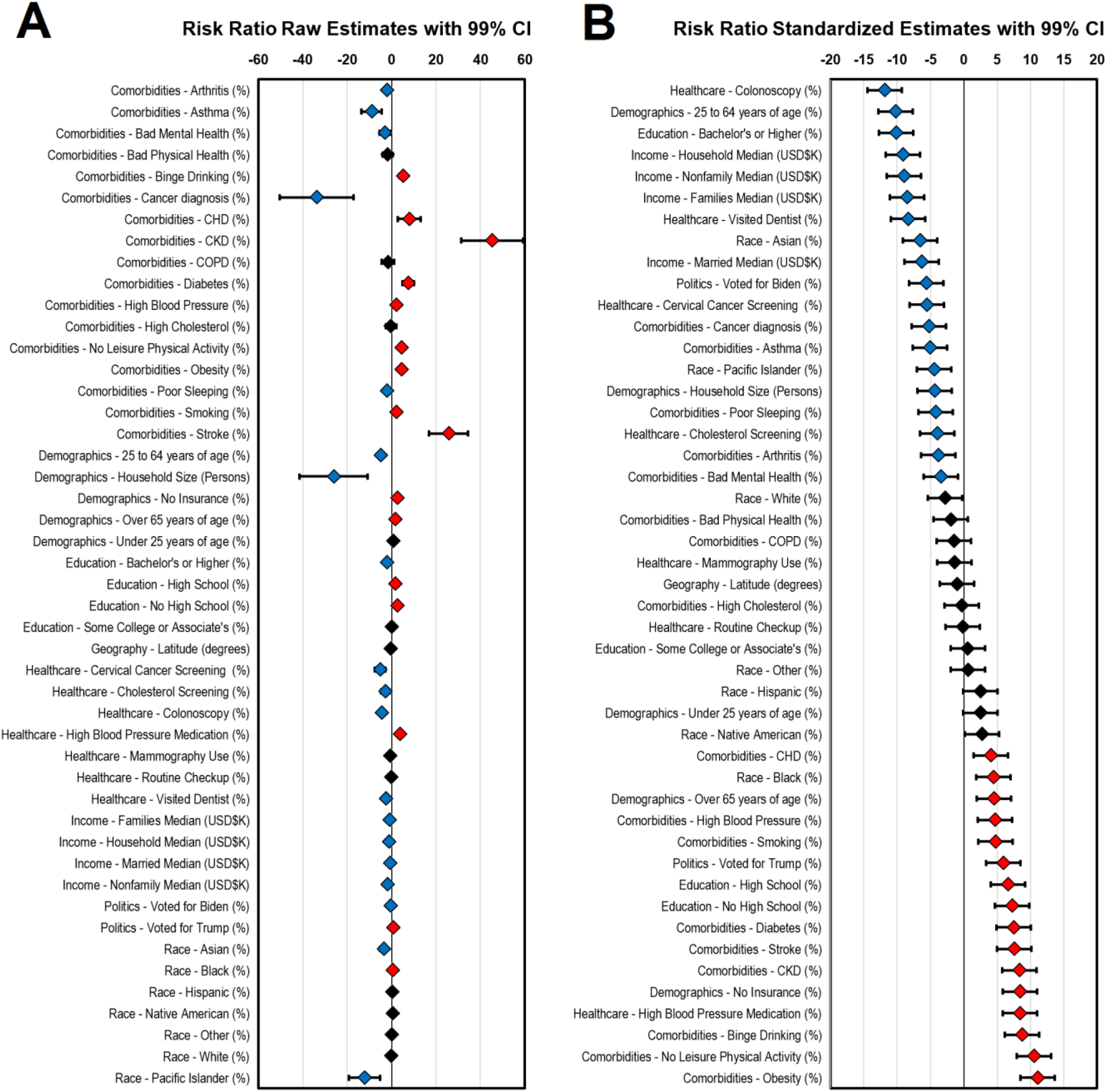
Comorbidity, sociodemographic and determinants of health associations to COVID-19 case-fatalities. All models are CCVI adjusted. A) Risk ratio raw estimates B) Risk ration standardized estimates (ordered). Blue diamonds indicate case-fatality reduction, red diamonds indicate case-fatality increase, and Black diamonds indicate no association. A total of 3140 counties were included in the study. Even when 99% CI are presented, association are declared significant at a Bonferroni adjusted threshold (47 tests P_adj_≤1.06E-03).

### Regional trends

The main analysis was also replicated independently within each of the ten US Health and Human Services defined regions. These models were also adjusted by CCVI. Risk ratio effect estimates for the ten regions are displayed in Figure 2. These analyses detected a wide array of effects that in some cases go in opposite directions across all regions. No single factor was consistently associated for all regions suggesting that regional associations are not generalizable. These regional assessments all have different sample sizes that are based on the number of counties within each state. These can range from 67 (Region 1) to 736 (Region 4); however, this discrepancy did not affect the capacity of each regional analysis to detect associations at a Bonferroni level (adjusted for 470 tests across all sets). The top variables reducing and increasing COVID-19 case-fatalities for each region are presented in Figure 3. The map in Figure 3A shows that the strongest protective regional effects were observed towards the east of the country where Stroke and Cancer Diagnosis were highly protective in the Northeast regions (Regions 1 and 2) and being of Pacific Islander descent was protective in the Southeast United States (Region 4). The Midwest displayed some moderate protective effects where household size was the top reducing factor in two regions (Region 5 and 7). Western regions displayed smaller COVID-19 case-fatality protective effects. Regions in the Western United States displayed smaller effect sizes in comparisons to regions in the South or Midwest. Lastly, Figure 3B shows variables that most significantly contributed to increased COVID-19 case-fatalities. Chronic kidney disease (CKD) was the most prevalent comorbidity across several regions (Regions 4, 6, 8 and 9). Among other findings, household size had a negative impact on COVID-19 outcomes specifically in the Northeast regions (Regions 1 and 2). As previously mentioned, some of the top variables displayed opposing effects which suggested that the interpretation must be done in context with the specific characteristics of that region, and interpretations cannot be generalized to others.

**Figure 2.**
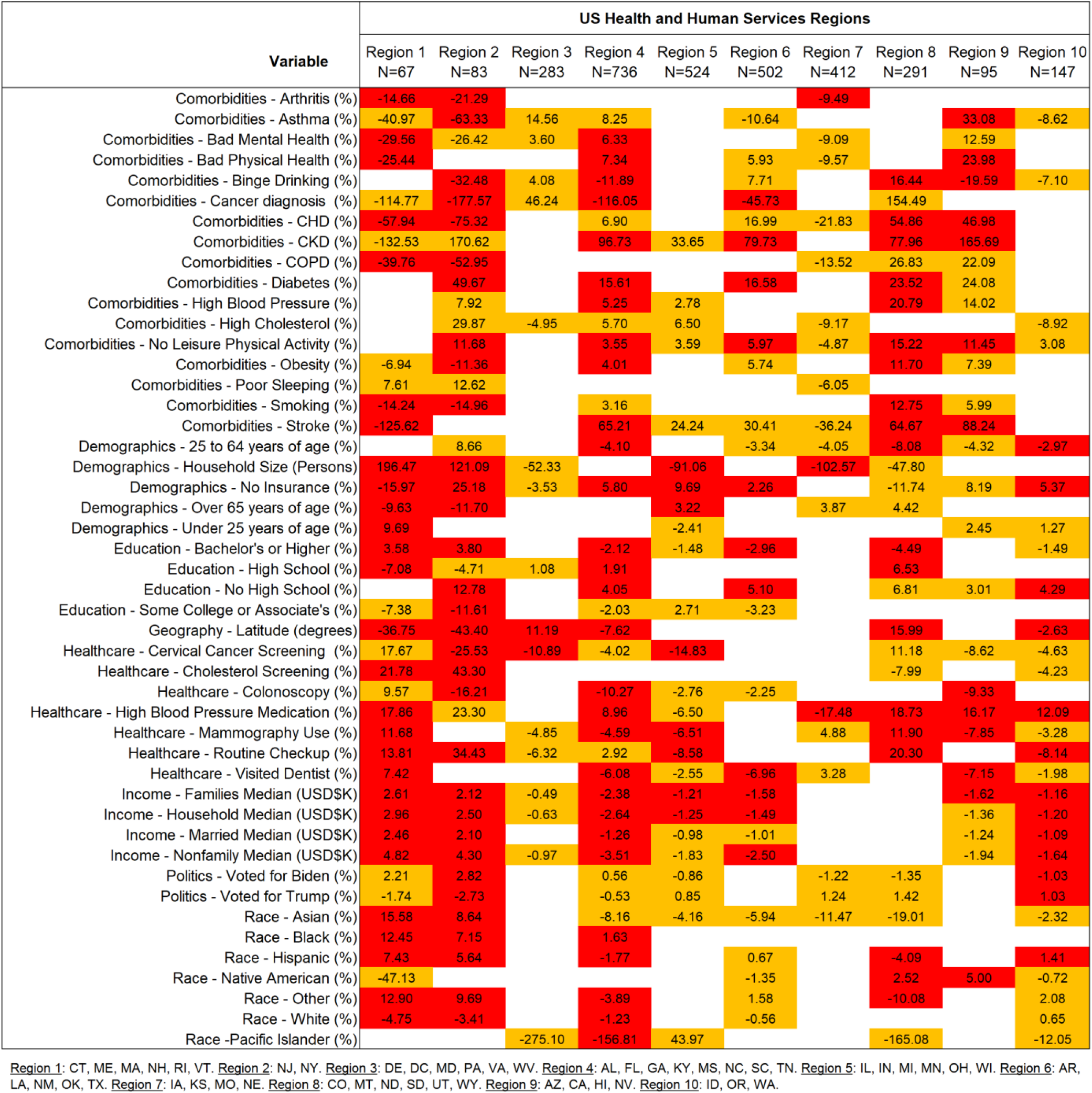
Regional risk ratio estimates for comorbidity, sociodemographic and determinants of health in association to COVID-19 case-fatalities. All models were performed independently by region and are adjusted to CCVI. Sample sizes correspond to the number of counties in the analysis for each region. Estimates signs indicate effect direction. Red boxes are significant to a Bonferroni level (470 tests P_adj_≤1.06E-04). Orange boxes are significant to a 95% confidence level.

**Figure 3.**
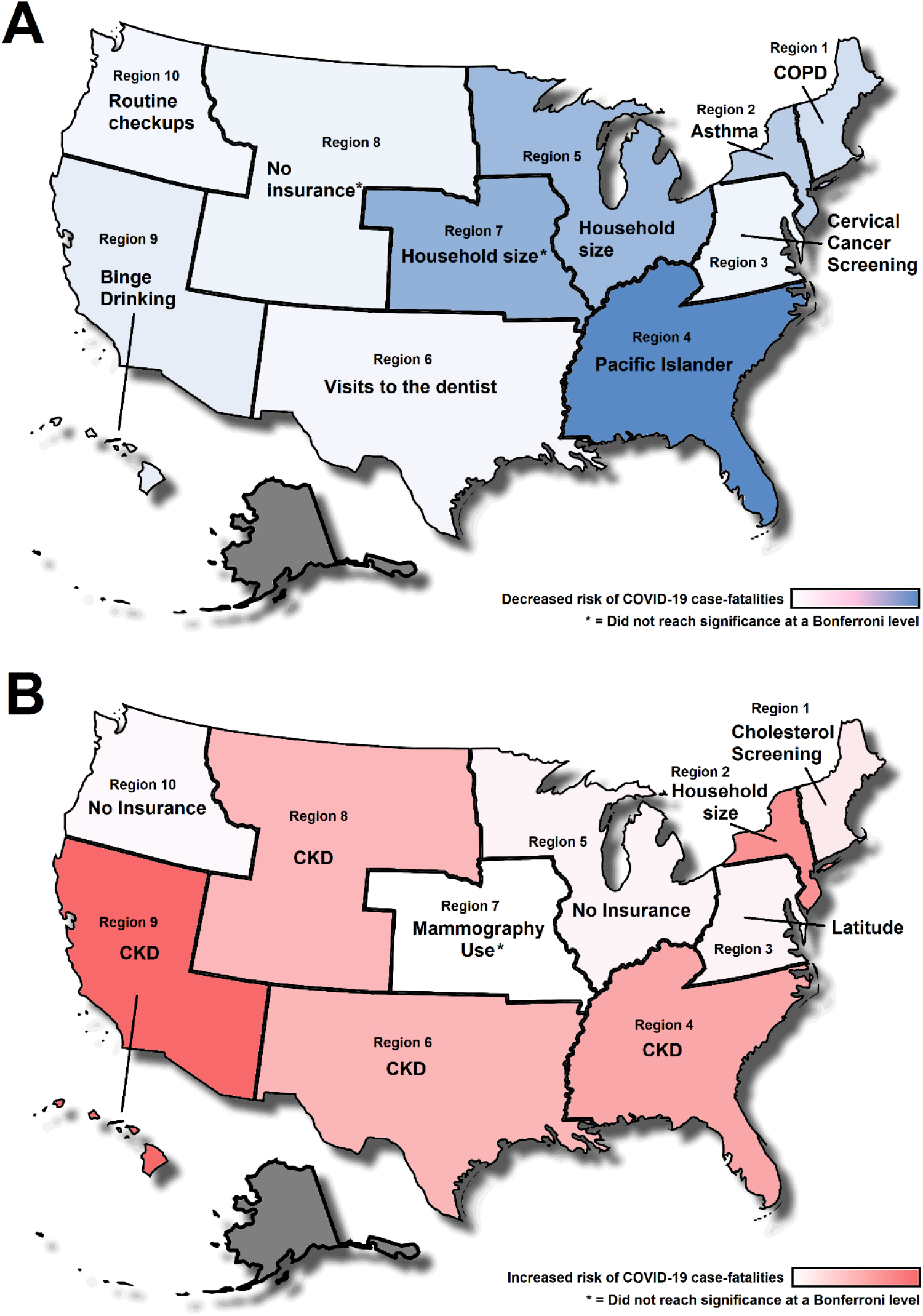
Top county level factors associated to COVID-19 case-fatalities for each United States Department of Health and Human Services regions. A) Top factors associated with reduced case fatalities B) Top factors associated with increased case-fatalities. Shading indicates the effect size across regions (adjusted by CCVI). Alaska was excluded from the analysis because of differences in their county level reporting

## DISCUSSION

The objective of this paper was to evaluate COVID-19 case-fatality rate in conjunction with sociodemographic factors, comorbidities, and determinants of heath. Previous studies have evaluated the influence of various socioeconomic factors ^7,14,24^ and comorbidities ^4–6^ on COVID-19 case-fatality rate; however, these analyses do not pair together their findings to be comparable with each other. Our study evaluates COVID-19 fatality rates from a wide timeframe, without potential influence from vaccines reducing case-fatality rate and the addition of major COVID-19 variants. Our study builds on the efforts of previous studies by presenting together a wide array of variables that describe community characteristics. It is necessary to emphasize that the associations between these factors and case-fatalities is not necessarily or entirely causative. All comorbidities, sociodemographic and determinants of health variables presented describe characteristics of the population that are not isolated or independent. Therefore, the causality that could be attributed to each factor evaluated must always be provided with context as a community indicator as they are all dependent or interconnected on each other, examples of this are binge drinking, mammography and visits to the dentist rates, which are likely indirectly describing a characteristic of the community. In general, all these variables must be interpreted in a continuum of causality that can vary across regions depending on the context.

Chronic kidney disease rates were the strongest predictor of increase COVID-19 case-fatality in several US regions. This relationship is likely to be predominantly causal due to people with this condition being medically vulnerable ^25–28^. Other comorbidities followed a similar trend such as higher rates of hypertension, diabetes, stroke, and smoking being associated with an increased case-fatality rate in a specific county. This finding aligns with other studies ^5,29,30^ linking increased rates of comorbidities to poorer COVID-19 outcomes. Even though comorbidities were most often associated with worse COVID-19 outcomes, stroke and cancer diagnosis were linked to reduced case-fatality rates in the Northeast region. We speculate that a potential explanation for this relationship is possible more frequent mask usage ^31^ and more precautions taken by this group of people ^32^. Northeastern states had mask adherence rates greater than 75% during the latter part of 2020 ^31^ with usage potentially diminishing the influence of comorbidities such asthma and COPD on COVID-19 case-fatality rate. Those with asthma and COPD in these communities were maybe more likely to wear a mask, further reducing their chance of acquiring and succumbing to COVID-19.

Household size was identified early on to be a risk factor for COVID-19 transmission ^33^. Our results have shown a contrasting effect that supports this notion in the Northeast but has the opposite effect in the Midwest. The difference between these regions is likely related to the specific context of living conditions. Although the mean household size for regions 1 and 2 is not far from the mean household size for regions 5 and 7, with 2.48 and 2.41 respectively, this difference may capture differences in housing quality and composition ^34^. This may also be indirectly identifying behavioral factors that are not obvious but can be implied such as house proximity (higher in cities) which can affect the capacity of self-isolation. Population density could be partially be influenced by household size, which has shown to have an impact on transmission ^35^. The Northeast states have higher population densities when compared to midwestern states. In summary, living conditions, housing quality and composition, and population density all be important component that define the impact of household size on case fatality rate. Generally, higher income was also associated with decreased COVID-19 case fatality rates. There could be a multitude of reasons why a higher income may be beneficial. This could include not being classified as an essential worker leading to being more likely to take time off or work from home ^36^, or even being able to live outside high-density population and compact housing areas ^7,37^. Income is often reflective and associated with racial discrepancies in COVID-19 outcomes ^24,38,39^. In a study examining neighborhood median income and COVID-19, when examining the neighborhoods, Black populations more often lived in neighborhoods with a significantly lower median income ($35,000) whereas White populations more often lived in wealthier neighborhoods ($63,000). Communities with lower median incomes are more often Medicaid patients and have COVID-19 complications that require invasive tactics such as mechanical ventilation ^40^. Income inequalities have been strong predictors of higher case numbers and fatalities throughout this pandemic ^14^.

The context that defines the influence of social dynamics on COVID-19 is complicated. Political affiliation has been repeatedly evaluated as a potential factor influencing the pandemic’s mortality ^41,42^. This pattern is not new, as Republican party affiliation was associated with decreased influenza vaccinations and the Democrat party was associated with increased compliance ^43,44^. The politicization of pandemic response has continued into the COVID-19 pandemic, with behaviors such as masking, social distancing, and vaccination being often divided along party lines. Some studies have shown a decrease in pandemic preventive health measures among Republicans while there has been increased adherence to public health recommendations among Democrats ^45,46^. Similarly, Republicans have been shown to have lower COVID-19 risk perception ^47,48^, when compared to other political parties which may influence their likelihood of contracting COVID-19. The behaviors exhibited by each political party may influence the results of this study. Our study showed that voting for Joseph Biden in the 2020 presidential election was mildly associated with decreased case-fatality rate while voting for Donald Trump was associated with increased rate. With Democrats being shown to be more likely to adhere to public health guidelines, they may be less likely to acquire and perish from COVID-19 while the inverse is true for Republicans. In December 2020, states with Republican governors had higher rates of cases, deaths, and positive tests than states with Democrat governors ^49^. This trend is evident in a similar approach using national data presented by NPR in collaboration with researchers at John’s Hopkins University where it was shown that voting Republican also had a deleterious effect ^50^. Rural and urban differences have been shown to play a major role in case-fatality rate as well with rural counties having a higher case-fatality ratio than urban counties ^4^. Rural voters are more likely to vote Republican ^51^andtherefore, the influence of politics in our findings may also be capturing geographical differences. Rural areas tend to have worse health outcomes in general and have significantly less access to care compared to their urban counterparts ^52^ These disparities add to the likelihood of developing comorbidities and ultimately, poorer COVID-19 outcomes. The link between political affiliation and COVID-19 case fatality rate is far more complex than the individual candidates that people of a county voted for. Political affiliation in our study is an indicator of underlying sociodemographic, health, and psychological trends that are more causative rather than associative.

### Limitations and future directions

Our study utilized aggregate data on a per-county basis instead of individual patient data; therefore, it is not possible to evaluate factors that contribute to COVID-19 case-fatality on a per case fashion which could help avoid any erroneous generalizations of specific regions. Another limitation of using county level data is that there is significant variability in the size and number of counties across the United States. Some counties may have only a few hundred people, while other counties may have a few million and this may lead, to some extent, representation bias. Future directions of this study include using these results to guide the evaluation of individuals’ factors that contribute directly to illness outcomes.

### Conclusion

Our study evaluated a multitude of factors that may affect COVID-19 case-fatality rate. Unlike previous studies that evaluated these factors separately, we performed a comprehensive analysis of all these variables together where they interact amongst each other. We identified several unique regionally dependent and independent relationships that highlighted the various factors that might influence COVID-19. Like other studies, we determined that comorbidities and demographic factors together are strong drivers of COVID-19 case fatalities. However, our study presents an assessment that puts them side to side for direct comparison. Our study highlights how any association is often dependent on the regional context. For example, household size in the Northeastern region of the United States was associated with more fatalities, while larger household size in the Midwest regions had a protective effect. Political voting patterns were also indicative of underlying healthcare patterns, with overall reduced case fatality rates in Democratic voting counties compared to increased fatality rates in Republican voting counties. The trends we identified in our study emphasize the importance of interpreting each factor in the context of other variables instead of in isolation.

## Data Availability

All data used in the study is publicly available. Curated datasets can be made available with a reasonable request to the corresponding author.

https://healthdata.gov/Health/COVID-19-Community-Profile-Report/gqxm-d9w9

https://chronicdata.cdc.gov/500-Cities-Places/PLACES-Local-Data-for-Better-Health-County-Data-20/swc5-untb/data

https://covid19.census.gov/datasets/USCensus::average-household-size-and-population-density-county/explore?location=3.973504%2C0.315550%2C2.69&showTable=true

https://www.census.gov/geographies/reference-files/time-series/geo/gazetteer-files.html

doi:doi:10.7910/DVN/VOQCHQ

https://www.census.gov/data.html

https://data.census.gov/cedsci/

## AUTHOR CONTIRBUTIONS

*Jacob Gerken*: Conceptualization, Methodology, Data Curation, Investigation, Writing of Original Draft. *Demi Zapata and Daniel Kuivinen*: Conceptualization, Methodology, Investigation, Writing of Original Draft. *Isain Zapata*: Conceptualization, Methodology, Investigation, Formal Analysis, Visualization, Supervision, Writing and Editing of Final Draft. All authors approved the final manuscript as submitted and agree to be accountable for all aspects of the work.

## REFERENCES

1. US Center for Disease Control. CDC COVID Data Tracker. Published 2022. Accessed April 4, 2022. https://covid.cdc.gov/covid-data-tracker/#cases_deathsper100k

2. Thorbecke W. The Impact of the COVID-19 Pandemic on the U.S. Economy: Evidence from the Stock Market. J Risk Financ Manag. 2020;13(10):233. doi:10.3390/jrfm13100233

3. Mascha EJ, Schober P, Schefold JC, Stueber F, Luedi MM. Staffing With Disease-Based Epidemiologic Indices May Reduce Shortage of Intensive Care Unit Staff During the COVID-19 Pandemic. Anesth Analg. 2020;131(1):24–30. doi:10.1213/ANE.0000000000004849

4. Iyanda AE, Boakye KA, Lu Y, Oppong JR. Racial/Ethnic Heterogeneity and Rural-Urban Disparity of COVID-19 Case Fatality Ratio in the USA: a Negative Binomial and GIS-Based Analysis. J Racial Ethn Heal Disparities. 2022;9(2):708–721. doi:10.1007/s40615-021-01006-7

5. Sarmadi M, Moghanddam VK, Dickerson AS, Martelletti L. Association of COVID-19 distribution with air quality, sociodemographic factors, and comorbidities: an ecological study of US states. Air Qual Atmos Heal. 2021;14(4):455–465. doi:10.1007/s11869-020-00949-w

6. Hashim MJ, Alsuwaidi AR, Khan G. Population Risk Factors for COVID-19 Mortality in 93 Countries. J Epidemiol Glob Health. 2020;10(3):204. doi:10.2991/jegh.k.200721.001

7. Karmakar M, Lantz PM, Tipirneni R. Association of Social and Demographic Factors With COVID-19 Incidence and Death Rates in the US. JAMA Netw Open. 2021;4(1):e2036462. doi:10.1001/jamanetworkopen.2020.36462

8. Peng M, He J, Xue Y, Yang X, Liu S, Gong Z. Role of Hypertension on the Severity of COVID-19: A Review. J Cardiovasc Pharmacol. 2021;78(5):e648–e655. doi:10.1097/FJC.0000000000001116

9. Wu Z, Tang Y, Cheng Q. Diabetes increases the mortality of patients with COVID-19: a meta-analysis. Acta Diabetol. 2021;58(2):139–144. doi:10.1007/s00592-020-01546-0

10. Yang D, Xiao Y, Chen J, et al. COVID-19 and chronic renal disease: clinical characteristics and prognosis. QJM An Int J Med. 2020;113(11):799–805. doi:10.1093/qjmed/hcaa258

11. Singh D, Mathioudakis AG, Higham A. Chronic obstructive pulmonary disease and COVID-19: interrelationships. Curr Opin Pulm Med. 2022;28(2):76–83. doi:10.1097/MCP.0000000000000834

12. Chung MK, Zidar DA, Bristow MR, et al. COVID-19 and Cardiovascular Disease. Circ Res. 2021;128(8):1214–1236. doi:10.1161/CIRCRESAHA.121.317997

13. Snowden LR, Graaf G. COVID-19, Social Determinants Past, Present, and Future, and African Americans’ Health. J Racial Ethn Heal Disparities. 2021;8(1):12–20. doi:10.1007/s40615-020-00923-3

14. Tan AX, Hinman JA, Abdel Magid HS, Nelson LM, Odden MC. Association Between Income Inequality and County-Level COVID-19 Cases and Deaths in the US. JAMA Netw Open. 2021;4(5):e218799. doi:10.1001/jamanetworkopen.2021.8799

15. White House COVID-19 Team. COVID-19 Community Profile Report. Accessed March 31, 2022. https://healthdata.gov/Health/COVID-19-Community-Profile-Report/gqxm-d9w9

16. Baer J, Campigotto C, Coome L, et al. Vulnerable Communities and COVID-19: The Damage Done, and the Way Forward. Surgo Ventur. Published online 2021.

17. US Center for Disease Control. 2020 PLACES. Local data for better health county data 2020 release. Published 2021. Accessed March 12, 2021. https://chronicdata.cdc.gov/500-Cities-Places/PLACES-Local-Data-for-Better-Health-County-Data-20/swc5-untb/data

18. US Census Bureau. Average Household Size and Population Density - County. Published 2020. https://covid19.census.gov/datasets/USCensus::average-household-size-and-population-density-county/explore?location=3.973504%2C0.315550%2C2.69&showTable=true

19. US Census Bureau. Gazetteer Files. Published 2021. https://www.census.gov/geographies/reference-files/time-series/geo/gazetteer-files.html

20. Lab MITED and S. County Presidential Election Returns 2000-2020. doi:doi:10.7910/DVN/VOQCHQ

21. US Census Bureau. 2020 Decennial United States Census. Published 2021. https://www.census.gov/data.html

22. US Census Bureau. 2019 American community survey 5-year public use microdata samples. Published 2020. Accessed March 12, 2021. https://data.census.gov/cedsci/

23. Melvin SC, Wiggins C, Burse N, Thompson E, Monger M. The Role of Public Health in COVID-19 Emergency Response Efforts From a Rural Health Perspective. Prev Chronic Dis. 2020;17:E70–E70. doi:10.5888/pcd17.200256

24. Liao TF, De Maio F. Association of Social and Economic Inequality With Coronavirus Disease 2019 Incidence and Mortality Across US Counties. JAMA Netw Open. 2021;4(1):e2034578. doi:10.1001/jamanetworkopen.2020.34578

25. Ortiz A, Cozzolino M, Fliser D, et al. Chronic kidney disease is a key risk factor for severe COVID-19: a call to action by the ERA-EDTA. Nephrol Dial Transplant. 2021;36(1):87–94. doi:10.1093/ndt/gfaa314

26. Jdiaa SS, Mansour R, El Alayli A, Gautam A, Thomas P, Mustafa RA. COVID–19 and chronic kidney disease: an updated overview of reviews. J Nephrol. 2022;35(1):69–85. doi:10.1007/s40620-021-01206-8

27. Flythe JE, Assimon MM, Tugman MJ, et al. Characteristics and Outcomes of Individuals With Pre-existing Kidney Disease and COVID-19 Admitted to Intensive Care Units in the United States. Am J Kidney Dis. 2021;77(2):190-203.e1. doi:10.1053/j.ajkd.2020.09.003

28. Gao Y, Ding M, Dong X, et al. Risk factors for severe and critically ill COVID-19 patients: A review. Allergy. 2021;76(2):428–455. doi:10.1111/all.14657

29. Yang J, Zheng Y, Gou X, et al. Prevalence of comorbidities and its effects in patients infected with SARS-CoV-2: a systematic review and meta-analysis. Int J Infect Dis. 2020;94:91–95. doi:10.1016/j.ijid.2020.03.017

30. Ejaz H, Alsrhani A, Zafar A, et al. COVID-19 and comorbidities: Deleterious impact on infected patients. J Infect Public Health. 2020;13(12):1833–1839. doi:10.1016/j.jiph.2020.07.014

31. Fischer CB, Adrien N, Silguero JJ, Hopper JJ, Chowdhury AI, Werler MM. Mask adherence and rate of COVID-19 across the United States. PLoS One. 2021;16(4):e0249891. https://doi.org/10.1371/journal.pone.0249891

32. Faria N, Costa MI, Gomes J, Sucena M. Reduction of Severe Exacerbations of COPD during COVID-19 Pandemic in Portugal: A Protective Role of Face Masks? COPD J Chronic Obstr Pulm Dis. 2021;18(2):226–230. doi:10.1080/15412555.2021.1904387

33. Madewell ZJ, Yang Y, Longini IM, Halloran ME, Dean NE. Household Transmission of SARS-CoV-2. JAMA Netw Open. 2020;3(12):e2031756. doi:10.1001/jamanetworkopen.2020.31756

34. Brown CC, Young SG, Pro GC. COVID-19 vaccination rates vary by community vulnerability: A county-level analysis. Vaccine. 2021;39(31):4245–4249. doi:https://doi.org/10.1016/j.vaccine.2021.06.038

35. Sy KTL, White LF, Nichols BE. Population density and basic reproductive number of COVID-19 across United States counties. Ndeffo Mbah ML, ed. PLoS One. 2021;16(4):e0249271. doi:10.1371/journal.pone.0249271

36. McCormack G, Avery C, Spitzer AK-L, Chandra A. Economic Vulnerability of Households With Essential Workers. JAMA. 2020;324(4):388. doi:10.1001/jama.2020.11366

37. Hamidi S, Hamidi I. Subway Ridership, Crowding, or Population Density: Determinants of COVID-19 Infection Rates in New York City. Am J Prev Med. 2021;60(5):614–620. doi:10.1016/j.amepre.2020.11.016

38. Escobar GJ, Adams AS, Liu VX, et al. Racial Disparities in COVID-19 Testing and Outcomes. Ann Intern Med. 2021;174(6):786–793. doi:10.7326/M20-6979

39. Mackey K, Ayers CK, Kondo KK, et al. Racial and Ethnic Disparities in COVID-19–Related Infections, Hospitalizations, and Deaths. Ann Intern Med. 2021;174(3):362–373. doi:10.7326/M20-6306

40. Quan D, Luna Wong L, Shallal A, et al. Impact of Race and Socioeconomic Status on Outcomes in Patients Hospitalized with COVID-19. J Gen Intern Med. 2021;36(5):1302–1309. doi:10.1007/s11606-020-06527-1

41. Gollwitzer A, Martel C, Brady WJ, et al. Partisan differences in physical distancing are linked to health outcomes during the COVID-19 pandemic. Nat Hum Behav. 2020;4(11):1186–1197. doi:10.1038/s41562-020-00977-7

42. Christopher W, Lynn V, Ryan B-K. Fatalities from COVID-19 are reducing Americans’ support for Republicans at every level of federal office. Sci Adv. 2022;6(44):eabd8564. doi:10.1126/sciadv.abd8564

43. Mesch GS, Schwirian KP. Social and political determinants of vaccine hesitancy: Lessons learned from the H1N1 pandemic of 2009-2010. Am J Infect Control. 2015;43(11):1161–1165. doi:10.1016/j.ajic.2015.06.031

44. Baum MA. Red State, Blue State, Flu State: Media Self-Selection and Partisan Gaps in Swine Flu Vaccinations. J Health Polit Policy Law. 2011;36(6):1021–1059. doi:10.1215/03616878-1460569

45. Rabin C, Dutra S. Predicting engagement in behaviors to reduce the spread of COVID-19: the roles of the health belief model and political party affiliation. Psychol Health Med. 2022;27(2):379–388. doi:10.1080/13548506.2021.1921229

46. Milligan MA, Hoyt DL, Gold AK, Hiserodt M, Otto MW. COVID-19 vaccine acceptance: influential roles of political party and religiosity. Psychol Health Med. Published online August 18, 2021:1-11. doi:10.1080/13548506.2021.1969026

47. Gratz KL, Richmond JR, Woods SE, et al. Adherence to Social Distancing Guidelines Throughout the COVID-19 Pandemic: The Roles of Pseudoscientific Beliefs, Trust, Political Party Affiliation, and Risk Perceptions. Ann Behav Med. 2021;55(5):399–412. doi:10.1093/abm/kaab024

48. Bilewicz M, Soral W. The Politics of Vaccine Hesitancy: An Ideological Dual-Process Approach. Soc Psychol Personal Sci. Published online November 15, 2021:19485506211055296. doi:10.1177/19485506211055295

49. Neelon B, Mutiso F, Mueller NT, Pearce JL, Benjamin-Neelon SE. Associations Between Governor Political Affiliation and COVID-19 Cases, Deaths, and Testing in the U.S. Am J Prev Med. 2021;61(1):115–119. doi:10.1016/j.amepre.2021.01.034

50. Wood D, Brumfiel G. Pro-Trump counties now have far higher COVID death rates. Misinformation is to blame. NPR. https://www.npr.org/sections/health-shots/2021/12/05/1059828993/data-vaccine-misinformation-trump-counties-covid-death-rate. Published December 5, 2021.

51. Parker K, Horowitz J, Brown A, Fry R, Cohn D, Igielnik R. Urban, suburban and rural residents’ views on key social and political issues. Pew Res Cent. Published online 2018.

52. James C V., Moonesinghe R, Wilson-Frederick SM, Hall JE, Penman-Aguilar A, Bouye K. Racial/Ethnic Health Disparities Among Rural Adults — United States, 2012–2015. MMWR Surveill Summ. 2017;66(23):1–9. doi:10.15585/mmwr.ss6623a1

